# Genome-wide scan of Flortaucipir PET levels finds *JARID2* associated with cerebral tau deposition

**DOI:** 10.1101/2024.10.04.24314853

**Authors:** Tamil Iniyan Gunasekaran, Devendra Meena, Annie J Lee, Siwei Wu, Logan Dumitrescu, Reisa Sperling, Timothy J Hohman, Alzheimer’s Disease Neuroimaging Initiative and the A4 Study, Jingxian Huang, Abbas Dehghan, Ioanna Tzoulaki, Richard Mayeux, Badri Vardarajan

## Abstract

**BACKGROUND:** Genetic research on Alzheimer’s disease (AD) has primarily focused on amyloid-β (Aβ) pathogenesis, with fewer studies exploring tau pathology. Elucidating the genetic basis of tau pathology could identify novel pathways in AD.

**METHODS:** We conducted a genome-wide association study of tau standard uptake value ratios (SUVRs) from ^[18]^F-flortaucipir positron emission tomography (PET) images to identify genetic variants underlying Tau pathology. Genetic data and tau-SUVRs from ^[18]^F-flortaucipir PET images were acquired from the A4 (311 with preclinical AD) and ADNI (280 cognitively normal, 76 with mild cognitive impairment, and 19 AD patients) studies. Circulating plasma proteins in UK Biobank Pharma Proteomics Project (UKBPPP, N=54,129) were used to validate genetic findings. SNP genotypes were tested for association with Tau-SUVR levels adjusting for age, sex and population substructure variables. AD association of polygenic risk scores (PRS) of tau and amyloid-SUVRs were assessed. Causal effect of plasma protein levels on Tau pathology were tested using Mendelian randomization analyses.

**RESULTS:** GWAS of tau-SUVR revealed two significant loci: rs78636169 (*P*=5.76×10^-10^) in *JARID2* and rs7292124 (*P*=2.20×10^-8^) near *ISX*. Gene-based analysis of tau deposition highlighted *APOE* (*P*=2.55×10^-6^), *CTNNA3* (*P*=2.86×10^-6^) and *JARID2* (*P*=1.23×10^-4^), a component of the *PRC2* multi-protein complex which regulates gene expression. Mendelian randomization analysis of available circulating plasma proteins in the UK Biobank Pharma Proteomics Project (UKBPPP) identified LRRFIP1, a protein that binds with *PRC2* multi-protein complex, as potentially causally linked to tau pathology. Genes associated with both amyloid and tau pathologies were enriched in endocytosis and signal transduction pathways. AD polygenic risk score (PRS) was associated with amyloid-SUVR but not with tau-SUVR. Amyloid-SUVR PRS had a notable association with AD clinical status, particularly in younger *APOE*-ε4 carriers, whereas tau-SUVR PRS showed a stronger association in older carriers.

**CONCLUSION:** We identified a novel potential therapeutic target, *JARID2* in the *PRC2* multi-protein complex, for tau pathology. Furthermore, gene pathway analysis clarified the distinct roles of Aβ and tau in AD progression, underscoring the complexity of genetic influences across different stages of the disease.

## Introduction

Alzheimer’s disease (AD) is a complex neurodegenerative disease characterized by the abnormal deposition of extracellular amyloid-β (Aβ) protein in neuritic plaques and intracellular hyperphosphorylated tau protein in neurofibrillary tangles within the brain ^1^. Neurodegeneration in AD is accompanied by hyperphosphorylated forms of tau. Cognitive impairment begins when tau accumulation spreads from the medial temporal lobe to the neocortex ^2^. Despite the crucial role tau pathology plays in AD, the genetic risk factors and gene pathways associated with tau pathologies remain unclear.

Genome-wide association studies (GWAS) of cerebral Aβ measures acquired from positron emission tomography (PET) images have identified several genetic loci, including *APOE* ^3,4^*, BCHE* ^4^*, IL1RAP* ^5^*, CR1, ABCA7,* and *FERMT2* ^3^. In our previous study, we conducted a large multi-ethnic meta-analysis of amyloid PET measures and identified a novel loci in the *RBFOX1* gene ^6^. Unlike amyloid PET images, tau PET scans are available in limited numbers due to the significant delay in the approval of tau tracers ^7^. Studies combining amyloid PET, tau PET, and structural magnetic resonance imaging (MRI) have identified tau accumulation as a primary contributor to cognitive decline in AD. This underscores the importance of tau PET imaging in evaluating AD-associated cognitive decline and neurodegeneration ^8–10^.

In a candidate gene study, SNPs in *BIN1* were associated with tau deposition ^11^. Previous tau-PET GWAS studies have pinpointed a few genetic loci, including *PPP2R2B, IGF2BP3* ^12^*, ZBTB20,* and *EYA4* ^13^, associated with tau deposition in the brain. However, these tau PET GWASs were carried out using single cohorts with limited sample sizes.

In this study, we harmonized tau PET measures from two studies and conducted a meta-analysis to identify genetic loci associated with tau deposition in the brain. Subsequently, we performed Mendelian randomization analysis of circulating plasma proteins in UK Biobank to identify proteins whose cognate genes may have causal link to tau deposition. We hypothesized that common genetic pathways could drive amyloid and tau pathologies in AD progression. Therefore, we analyzed genes, and their enriched pathways associated with both amyloid and tau pathologies. We also evaluated the predictive accuracy of tau polygenic scores (PRS) and amyloid PRS for clinical and pathological AD status.

## Materials and Methods

### Study participants

We obtained data from the Anti-Amyloid Treatment in Asymptomatic Alzheimer’s Disease (A4) study and the Alzheimer’s Disease Neuroimaging Initiative (ADNI) study, which are available in the ADNI database (http://adni.loni.usc.edu). Our analysis included data from 330 pre-clinical individuals from the A4, together with 303 cognitively normal (CN) individuals, 81 individuals with mild cognitive impairment (MCI), and 19 individuals with diagnosed AD from the ADNI. In ADNI, the clinical diagnosis of probable AD was based on the National Institute of Neurological Disorders and Stroke/Alzheimer’s Disease and Related Disorders Association (NINDS-ADRDA) criteria ^14^. Eligibility criteria for participation in the ADNI study are described elsewhere ^14^. Detailed information about the A4 study participants can be found in our previous study ^6^. Demographic characteristics of the study participants are presented in **Table 1**.

**Table 1:** Demographic characteristic of multi-ethnic population and non-Hispanic Whites for Tau PET from ADNI and A4 cohorts.

| Demographic information | ADNI |  |  |  |  |  | A4 |  |
| --- | --- | --- | --- | --- | --- | --- | --- | --- |
|  | All ethnicity |  |  | Non-Hispanic White |  |  | All ethnicity | Non-Hispanic White |
| Clinically diagnosed sample groups | CN | MCI | AD | CN | MCI | AD | Pre-clinical AD | Pre-clinical AD |
| N | 303 | 81 | 19 | 280 | 76 | 19 | 330 | 311 |
| Age (y, mean ± SD) | 71.43 ± 5.94 | 69.96 ± 7.62 | 73.79 ± 10.75 | 71.64 ± 5.87 | 70.28 ± 7.61 | 73.79 ± 10.75 | 78.41 ± 4.76 | 71.76 ± 4.76 |
| Female (n, %) | 184 (61%) | 32 (39%) | 7 (37%) | 164 (59%) | 28 (37%) | 7 (37%) | 206 (62%) | 193 (62%) |
| Years of education (y, mean ± SD) | 16.81 ± 2.38 | 16.36 ± 2.60 | 16.16 ± 2.75 | 16.82 ± 2.35 | 16.41 ± 2.63 | 16.16 ± 2.75 | 16.34 ± 2.73 | 16.33 ± 2.72 |
| MMSE (mean ± SD) | 29.18 ± 1.08 | 28.26 ± 1.51 | 22.68 ± 2.31 | 29.18 ± 1.10 | 28.24 ± 1.51 | 22.68 ± 2.31 | 28.73 ± 1.31 | 28.74 ± 1.29 |
| Tau SUVR (mean ± SD) | 1.09 ± 0.07 | 1.14 ± 0.14 | 1.36 ± 0.33 | 1.10 ± 0.07 | 1.14 ± 0.14 | 1.36 ± 0.33 | 1.08 ± 0.06 | 1.08 ± 0.06 |

ADNI, Alzheimer's Disease Neuroimaging Initiative; A4, Anti-Amyloid Treatment in Asymptomatic Alzheimer's Disease; CN, Cognitively Normal; MCI, Mild Cognitive Impairment; AD, Alzheimer's disease; N, number of participants; y, Years; SD, Standard Deviation; MMSE Mini Mental State Examination; SUVR, standard uptake value ratio

### PET imaging and processing

The baseline preprocessed ^[18]^F-flortaucipir PET (tau PET) images of both A4 and ADNI studies were downloaded from the ADNI database (https://ida.loni.usc.edu/). The detailed acquisition procedures were previously reported ^6,15,16^. T1-weighted magnetic resonance images (MRIs) that were acquired closest to the date of the baseline tau PET image scans and the respective tau PET images from the same participants were normalized to the standard native space using the SPM12 toolbox implemented in MATLAB (R2022a, Mathworks, Natick, MA, USA). The MRI images were processed using FreeSurfer, version 7.3.2 (https://surfer.nmr.mgh.harvard.edu/) to create anatomical regions of interest (ROIs) within a native space. Then, the tau PET images were co-registered with the respective FreeSurfer processed MRI data, and the SUVRs were calculated from the composite regions normalized to the cerebellar gray matter region, as described in the ADNI methods (https://adni.bitbucket.io/reference/docs/UCBERKELEYAV1451/UCBERKELEY_AV1451_Methods_2021-01-14.pdf). Pre-processed amyloid-PET SUVR scores from the ADNI and A4 participants were accessed as described here^17^.

### GWAS methods and statistical analysis

Description of the SNP data and quality control measures for GWAS analysis are described in the Supplementary Methods and **Supplementary Table 1**. SNP-based GWAS was separately performed with Z-score standardized tau-SUVR measures adjusting for age, sex, and the first three principal components (PCs) in the A4 dataset and additionally AD diagnosis in the ADNI dataset. A second model including *APOE*-ε4 dosage as a covariate was also tested. The analyses were conducted using PLINK software, version 1.9 ^18^. Results from A4 and ADNI studies were summarized in an inverse-weighted meta-analysis using METAL software ^19^.

Genome-wide gene-based analysis of tau-SUVR was implemented independently in A4 and ADNI using the MAGMA tool, version 1.09a ^20^ with the ‘multi=all’ function, using the same covariates as described above. Results were summarized using the ‘meta’ function in MAGMA. Similarly, gene-based amyloid-SUVR analysis were conducted using summary statistics from a previous amyloid PET GWAS ^6^.

Manhattan and quintile-quintile (Q-Q) plots for the SNP-based and gene-based GWAS results were generated using the ’qqman’ package in the R program, version 4.3.1 ^21^. Pathways enrichment analyses was conducted in genes that were nominally significant with tau and amyloid SUVRs (Supplementary Methods). Replication analysis was performed using summary statistics from a previously published tau-PET GWAS ^12^.

### Mendelian Randomization analysis

We applied the Mendelian Randomization (MR) framework to investigate whether genetically predicted circulating levels of plasma proteins are causally linked with Tau PET levels using the GWAS summary data from A4 and ADNI.

### Study population and data sources (Genetic associations of circulating plasma proteins)

Genetic associations of circulating plasma proteins were obtained from the UK Biobank Pharma Proteomics Project (UKBPPP) for individuals of European ancestry (N=54,129; up to 95% Europeans) ^22^. We limited our analyses to circulating plasma proteins whose cognate gene showed suggestive significance in our gene-based test (*P*_gene-based test_<0.05). Details about the instrument selection and MR analysis are described in Supplementary Methods.

### Polygenic risk score (PRS) analysis

PRSs for clinical AD in the ADNI/A4 participants and tau and amyloid SUVRs were in the Religious Orders Study and the Memory and Aging Project (ROSMAP) dataset were calculated using the PRSice tool, version 2, and PLINK software. Details of the reference datasets, quality control and PRS methods are provided in Supplementary Methods and **Supplementary Table 2**.

### Association analysis of brain cortical atrophy

The effect of tau-associated SNPs on brain cortical atrophy was evaluated through general linear models (GLM) adjusting for age and sex, in MATLAB (R2022a, The Mathworks, Natick, MA, USA) using the Surfstat toolbox (http://www.math.mcgill.ca/keith/surfstat/).

## Results

### Tau-PET GWAS identifies the *JARID2* locus

The tau SUVR distribution in the ADNI cohort was right-skewed compared to that of A4 cohort (**Figure 1A-1D**). MCI and AD patients in ADNI had higher levels of tau-deposition compared to CN in ADNI and A4 participants, who had similar levels. We used clinical AD as a covariate for the ADNI dataset, while no such adjustments were made for A4.

**Figure 1.**
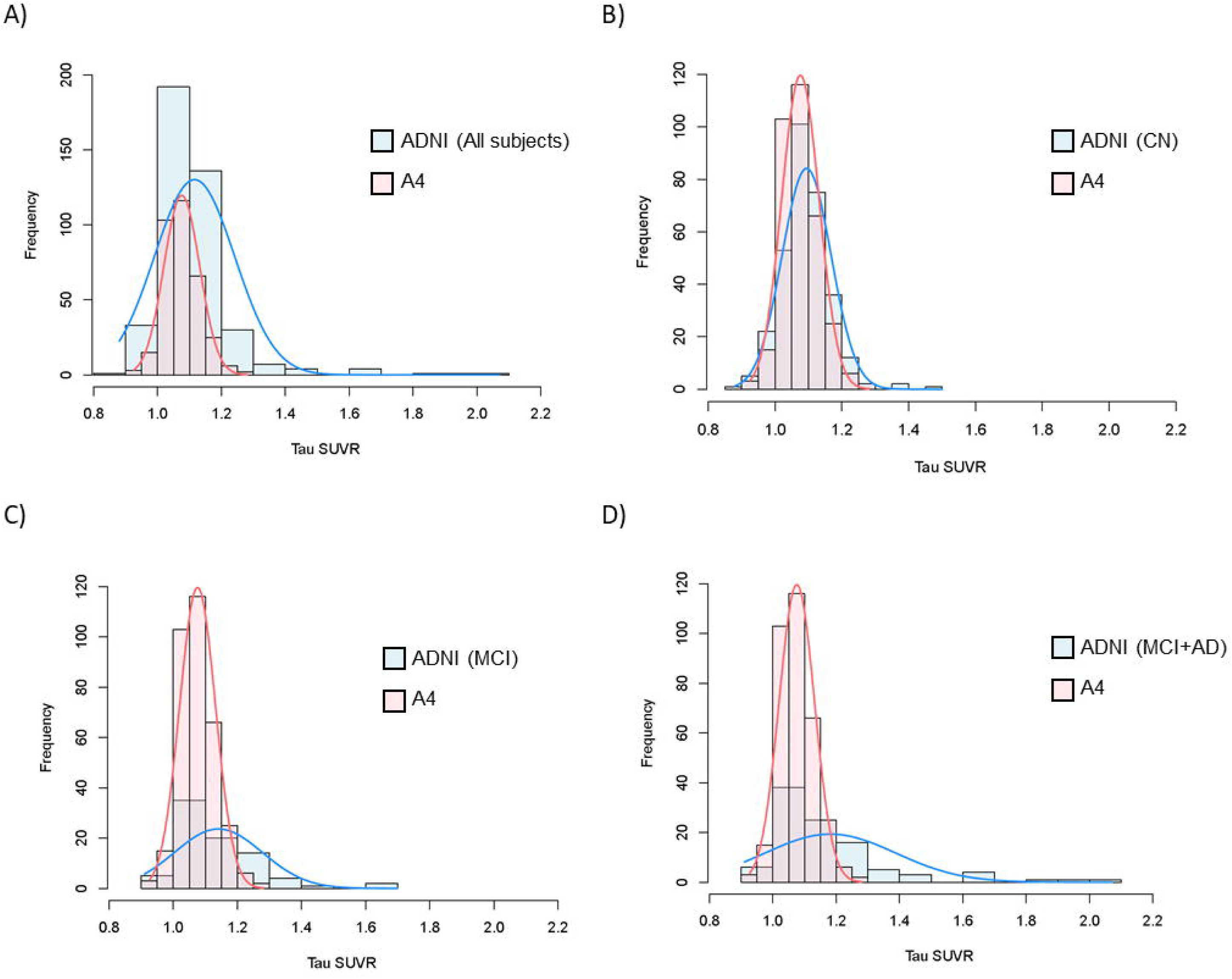
Tau standard uptake value ratios (SUVRs) histograms for ADNI and A4 cohort individuals. A) All individuals from both cohorts. B) Cognitively normal (CN) individuals from ADNI and all individuals from A4. C) Mildly cognitively impaired (MCI) individuals from ADNI and all individuals from A4. D) MCI and Alzheimer’s disease (AD) individuals from ADNI compared with all individuals from A4.

The SNP-based genome-wide meta-analysis of non-Hispanic White (NHW) individuals from A4 (N=311) and ADNI (N=375), identified a significant association of rs78636169 (*P*=5.76×10^-10^) near the *JARID2* (Jumonji and AT-Rich Interaction Domain containing 2) gene on chromosome 6 with tau deposition (**Figure 2A-2D**, **Table 2**, **and Supplementary Table 3**). The association remained genome-wide significant when we included all multi-ethnic individuals (**Supplementary Figure 1 and Supplementary Table 4**). In addition, rs7292124 (*P*=2.20×10^-8^), near the *ISX* (Intestine Specific Homeobox) gene on chromosome 22, was also genome-wide significant in NHW. These associations remained significant after adjustment for *APOE*-ε4 status, (rs78636169, *P*=1.20×10^-8^ and rs7292124, *P*=5.17×10^-9^). Additionally, rs112518363 (*P*=3.51×10^-8^) near *INTS10* (Integrator Complex Subunit 10) became genome-wide significant when adjusted for *APOE* ε4 status.

**Figure 2.**
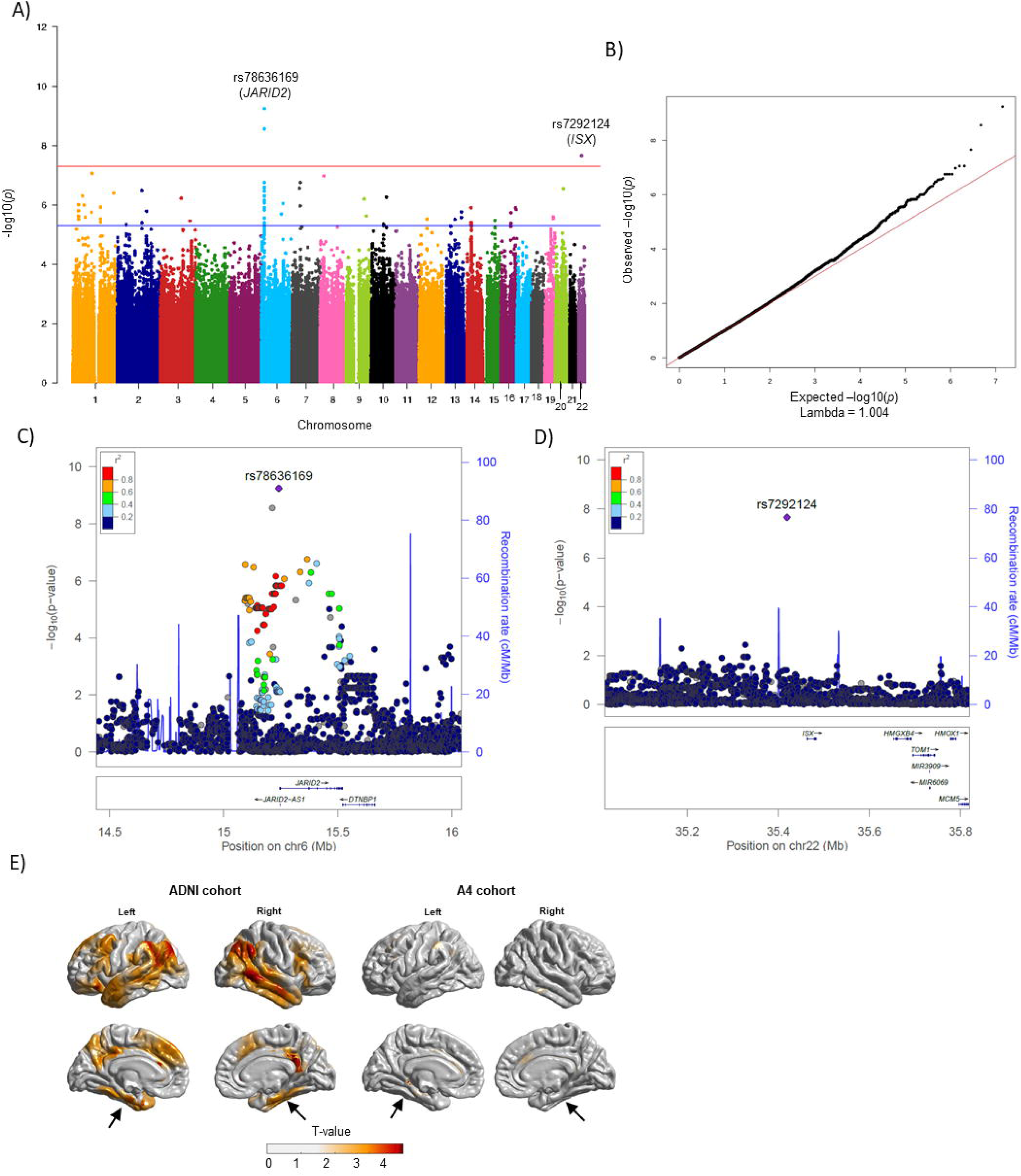
SNPs associated with cerebral tau deposition. (A) Manhattan plot showing meta-analysis *P*-values (depicted on the –log_10_ scale) from linear regression on cerebral tau deposition involving non-Hispanic White (NHW) subjects. The threshold for genome-wide significance is represented by a blue line at *P* = 5×10^-8^, while suggestive significance is indicated by a blue line at *P =* 5×10^-6^ threshold. (B) Quantile-Quantile (QQ) plots for the SNPs associated with cerebral tau deposition involving multi-ethnic subjects. The QQ plot showed no spurious genomic inflation (λ = 1.004). (C) Magnified LocusZoom regional association plots show the regions around rs78636169 in the *JARID2* loci and (D) rs7292124 near *ISX* loci. The purple dot indicates the most associated SNP in the region. E) Brain cortical atrophy associated with rs78636169 in *JARID2.* Brian cortical patterns associated with rs78636169 represented in t-value. rs78636169 AA+AG carriers were compared with GG carriers to infer the cortical atrophy in ADNI cohort (left) and A4 cohort (right). The black arrow indicates greater atrophy in the parahippocampal region among rs78636169 AA+AG carriers in the ADNI cohort, while mild atrophy was observed in the same region among individuals from the A4 cohort.

**Table 2.** Genome-wide significant associations with tau deposition among non-Hispanic White participants.

| SNP | Nearest gene | Chr | BP | Variant type | A1 | ADNI cohort |  |  |  | A4 cohort |  |  |  | Tau-SUVr Meta-analysis* |  |  |  |  |  |
| --- | --- | --- | --- | --- | --- | --- | --- | --- | --- | --- | --- | --- | --- | --- | --- | --- | --- | --- | --- |
| | | | | | | $\beta$ | SE | *P*-value | MAF | $\beta$ | SE | *P*-value | MAF | $\beta$ | SE | *P*-value | Dir | Het ChiSq | Het PVal |
| **rs78636169** | *JARID2* | 6 | 15242005 | Upstream variant | A | 0.73 | 0.17 | 1.66E-05 | 0.04 | 0.7 | 0.16 | 1.44E-05 | 0.06 | 0.71 | 0.12 | 5.76E-10 | ++ | 0.01 | 0.9 |
| **rs200751686** | *JARID2* | 6 | 15213555 | Intergenic region | CA | 0.66 | 0.16 | 6.70E-05 | 0.05 | 0.7 | 0.16 | 1.65E-05 | 0.06 | 0.68 | 0.11 | 2.74E-09 | ++ | 0.03 | 0.86 |
| **rs7292124** | *ISX* | 22 | 35419122 | Intergenic region | C | 0.77 | 0.16 | 1.75E-06 | 0.05 | 0.58 | 0.2 | 4.39E-03 | 0.04 | 0.69 | 0.12 | 2.20E-08 | ++ | 0.53 | 0.46 |

SNP, Single Nucleotide Polymorphism; Chr, Chromosome; BP, Base pair location; A1, Effect allele; SE, Standard error; MAF, Minor Allele Frequency; Dir, Effect direction; Het ChiSq, Chi-square value for heterogeneity test; Het PVal, *P*-value for heterogeneity in effect sizes in meta-analysis.
\**P*-value of 5E-08 was used to assess genome-wide significance in the meta-analysis.

**Table 3.** Mendelian Randomization identified proteins with potential causal effect on Tau PET levels.

| Mendelian Randomization analysis |  |  |  |  |  |  |  |  | Gene-based meta-analysis on non-Hispanic Whites showing the association with tau deposition |  |  |  |  |
| --- | --- | --- | --- | --- | --- | --- | --- | --- | --- | --- | --- | --- | --- |
| Protein | Method | Panel | NSNP | Beta | SE | OR (LCI-UCI) | P-value | FDR | GENE | Condition analysis 1* |  | Condition analysis 2<sup>#</sup> |  |
|  |  |  |  |  |  |  |  |  |  | ZSTAT | P-value | ZSTAT | P-value |
| LRRFIP1 | Inverse variance weighted | Inflammation II | 2 | -2.6353 | 0.7250 | 0.07 (0.02-0.3) | 2.78E-04 | 0.016 | *LRRFIP1* | 1.6247 | 0.05 | 2.6126 | 4.49E-03 |
| IST1 | Wald ratio | Inflammation II | 1 | -2.2590 | 0.6372 | 0.1 (0.03-0.36) | 3.93E-04 | 0.016 | *IST1* | 2.8723 | 2.04E-03 | 2.3256 | 0.01 |
| HADH | Wald ratio | Inflammation | 1 | 3.4662 | 1.0206 | 32.01 (4.33-236.62) | 6.83E-04 | 0.019 | *HADH* | 1.7522 | 0.04 | 2.741 | 3.06E-03 |
| PRTG | Inverse variance weighted | Oncology | 4 | -0.4152 | 0.1348 | 0.66 (0.51-0.86) | 2.07E-03 | 0.044 | *PRTG* | 1.5565 | 0.06 | 2.0335 | 0.02 |
| PRTG | Weighted median | Cardiometabolic | 4 | -0.3959 | 0.1413 | 0.67 (0.51-0.89) | 5.07E-03 | 0.044 |  |  |  |  |  |
| PRTG | MR Egger | Oncology | 4 | -0.5782 | 0.3128 | 0.56 (0.3-1.04) | 0.21 | 0.044 |  |  |  |  |  |

MR Egger, Mendelian Randomization-Egger; NSNP, Number of Single Nucleotide Polymorphism; SE, Standard Error; OR, Odds Ratio; LCI, Lower Confidence Interval; UCI, Upper Confidence Interval; FDR, False Discovery Rate; ZSTAT, Z statistics.
\*Condition analysis 1: Adjusted for age, sex, three principal components and clinical diagnosis status
<sup>#</sup>Condition analysis 2: Adjusted for age, sex, three principal components, *APOE* ε4 status and clinical diagnosis status

### Relationship of tau-associated SNPs with amyloid deposition and clinical Alzheimer’s Disease

Amongst 41,455 SNPs nominally associated (*P*<0.005) with tau deposition, we identified 20 genetic loci that are also associated (*P*<0.005) with amyloid-SUVR. Sixteen SNPs had the same direction of effect for both amyloid and tau deposition including SNPs related to *PVRL2* (Poliovirus receptor-related 2) and *APP* (Amyloid Precursor Protein) (**Supplementary Table 5**). Amongst 75 genetic loci associated for AD risk in a previous large-scale GWAS of clinical AD^23^, four variants in *SNORA73* (rs76928645), *INPP5D* (rs10933431), *ABCA7* (rs12151021) and *IL34* (rs4985556) were nominally associated (*P*<0.05) with tau pathology (**Supplementary Table 6**).

### Gene level association with tau deposition

Gene-based meta-analysis using MAGMA on tau-SUVR identified *APOE* (*P*=2.55×10^-6^) in chromosome 19 and *CTNNA3* (*P*=2.86×10^-6^) on chromosome 10 (**Supplementary Table 7**). *JARID2* had a suggestive association with tau deposition (*P*=1.22×10^-4^). Additionally, we conducted a gene-based analysis with amyloid deposition using summary statistics from our previous study^6^. In addition, we identified 61 genes that were nominally significant association (*P*<0.05) with both tau and amyloid deposition including *APOE*, *TOMM40*, and *COL5A2* (*P*<0.005) with both tau and amyloid pathologies (**Supplementary Figure 2 and Supplementary Table 8**). *JARID2* was also nominally significant (p=0.047) for association with Tau deposition in an independent tau-PET GWAS dataset ^12^ (**Supplementary Table 9)**.

### Mendelian randomization of circulating plasma proteins levels with tau deposition

Of the 916 genes nominally associated with tau SUVR levels, 84 overlapped with the UKBPPP and were used for the *cis*-MR analysis. Since JARID2 levels were not directly measured, we used other proteins in the PRC2 complex to confirm a causal link to Tau pathology. MR analysis of circulating plasma proteins found evidence of a potential causal effect of genetically predicted LRRFIP1 (β_Wald-ratio_ = -2.64 [-4.46, -1.21], p=0.016), HADH (β_Wald-ratio_ = 3.47 [1.47, 5.47], p=0.019), IST1 (β_Wald-ratio_ = -2.26 [-3.51, -1.01], p=0.016), and PRTG (β_IVW_ = -0.42 [-0.68, -0.15], p=0.04) on tau-PET levels (**Supplementary Figure 3, Table 4 and Supplementary Table 10**). Among these, HADH showed suggestive evidence of co-localization (PPH4=61.6%) while LRRFIP1 and IST1 indicated limited power in colocalization to differentiate between a true causal variant or horizontal pleiotropy (**Supplementary Figure 4 and Supplementary Table 11 and 12**).

### Pathways associated with tau and amyloid deposition

Pathway analysis revealed that genes associated with tau deposition were enriched in pathways related to the positive regulation of sterol and cholesterol transport, fascia adherens, postsynaptic specialization, and asymmetric synapse (*q*-value <0.05) (**Supplementary Figure 5 and Supplementary Table 13**).

Amongst genes showing significant associations with both tau and amyloid, no gene pathways remained significantly associated (*q*-value <0.05) after multiple corrections, but endocytosis and signal transduction pathways were nominally significant (**Supplementary Figure 6 and Supplementary Table 14**).

### Association of tau and amyloid PRS with clinical AD and neuropathological measures

In ADNI and A4 participants, we found a robust association of AD PRS with amyloid-SUVR levels (*P*=1.94×10^-11^; R^2^=0.079) but only a weak association with tau-SUVR (*P*=0.03; R^2^=0.0087).

Next, we analyzed the association of tau and amyloid-SUVR PRS with the clinical and pathological diagnosis of AD^24^ in the ROSMAP cohort. Results showed a significant association between amyloid PRS and clinical AD status (*P*=0.018; R^2^=0.0038), but not with tau PRS (*P*=0.29; R^2^=0.00076). However, tau PRS was significantly associated with clinical AD status among *APOE*-ε4 carriers (*P*=0.015; R^2^=0.017). In non-ε4 carriers, neither amyloid nor tau PRS were associated with clinical AD. Neither amyloid PRS nor tau PRS was associated with pathological AD status (**Supplementary Table 15**).

### Effects of rs78636169 on brain cortical atrophy

We analyzed the effects of the rs78636169 SNP from the *JARID2* gene on brain cortical atrophy under an additive genetic model. In ADNI, which included individuals with clinical diagnoses ranging CN, MCI and AD, the results revealed cortical atrophy patterns in the entorhinal cortex, parahippocampal region, posterior cingulate, inferior parietal lobe, precuneus, middle temporal gyrus, inferior temporal gyrus, and superior frontal region. Additionally, the atrophy pattern was asymmetrical in some regions **(Figure 2E**). Analysis of the A4 study cohort, which comprised pre-clinical individuals, showed cortical atrophy patterns in fewer regions, including parahippocampal, anterior cingulate and inferior parietal regions (**Figure 2E**).

## Discussion

In this study, we conducted a genome-wide meta-analysis using tau-SUVR measures derived from ^[18]^F-flortaucipir PET images from the A4 and ADNI study cohorts. Among 686 non-Hispanic White individuals, we identified two significant SNPs: rs78636169 (*P*=5.76×10^-10^) near *JARID2* and rs7292124 (*P*=2.20×10^-08^) near the *ISX* gene, associated with cerebral tau deposition. *JARID2* remained genome-wide significant when 47 multi-ethnic individuals were included and when adjusted for *APOE*-ε4 dosage, suggesting *APOE*-ε4 has no significant influence on these loci. Gene-based analyses identified *APOE* and *CTNNA3*, while *JARID2* showed suggestive significance. Our study reinforces the association of *APOE* with both amyloid tau deposition, ^4,6,25^.

*JARID2* (Jumonji, AT-rich interactive domain 2) is a protein coding gene involved in regulating gene expression and chromatin structure ^26^. JARID2 is also a component of PRC2 (Polycomb Repressive Complex 2) multi-protein complex, necessary for transcriptional silencing through histone modification^27^. PRC2 methylates JARID2, promoting PRC2 activity by guiding it to target genomic regions ^27,28^. PRC2 is essential for processes like cell differentiation, proliferation, and maintaining stem-cell plasticity. With age, *PRC2* expression decreases in the brain, leading to abnormal expression of genes linked to AD ^29^. Neuronal deficiency in *PRC2* is also associated with progressive neurodegeneration in mice ^30^. Overall, *JARID2* and *PRC2* multi-protein complex play roles in tau pathology and neurodegeneration in AD, though further validation is needed.

To establish causal relationship between circulating plasma protein levels of the top genetic hits, we conducted MR analysis using the UKBPPP cohort. Since JARID2 protein levels were not measured directly, we assessed constituent proteins in the PRC2 complex. MR analysis revealed potential causal links of genetically predicted LRRFIP1 protein levels to lower tau deposition. *LRRFIP1*, along with *PRC2* multi-protein complex, is involved in the downregulation of tumor necrosis factor-α (TNF-α) ^31^, a pro-inflammatory cytokine involved in regulating innate and adaptive immunity, playing an important role in AD pathology. Inhibiting TNF-α has demonstrated a protective effect against AD pathology, including Aβ and tau deposition ^32,33^. Our analysis suggests *JARID2, LRRFIP1* and other proteins in the *PRC2* multi-protein complex plays a critical role in protecting against tau pathology.

*CTNNA3* (α-3 catenin) forms the α-3/β-1 catenin complex with *CTNNB1* (β-1 catenin), binding to *PSEN1* (presenilin-1) and promoting higher Aβ42 levels ^34,35^, linked to the early onset of familial AD ^36,37^. *CTNNA3* is also associated with AD, particularly in females ^38^ and Caribbean-Hispanics^39^. In this study, *CTNNA3* shows an *APOE*-dependent association with tau deposition, consistent with previous studies ^40^.

We analyzed *MAPT* (Microtubule Associated Protein Tau), previously implicated in tau pathology in AD ^41,42^ and found that sixteen SNPs nominally associated with tau deposition, including rs63750072 (**Supplementary Table 16**), a missense mutation previously reported in AD-related studies ^43,44^.

SNPs from four AD-associated loci: *SNORA73, INPP5D, ABCA7,* and *IL34* showed nominal associations (*P*<0.05) with tau pathology. The roles of *ABCA7* (ATP Binding Cassette Subfamily A Member 7) ^45,46^, *IL34* (Interleukin 34) ^47,48^ and *INPP5D* (Inositol Polyphosphate-5-Phosphatase D) ^49,50^ in Aβ-mediated AD pathology have been well-studied. Our findings suggest a potential link between these genes and tau and amyloid pathologies.

Gene-based analysis from amyloid-PET and tau-PET GWASs identified 61 genes associated (*P*<0.05) with both amyloid and tau pathology. *APOE, TOMM40* and *COL5A2* showed robust associations (*P*<0.005). *COL5A2*, particularly noteworthy, is linked to reduced neuronal energy supply, leading to AD-related apoptosis ^51^. No previous studies have explored genes associated with both tau and amyloid pathologies using amyloid-PET and tau-PET GWAS results.

Gene pathway analysis revealed tau-associated genes enriched in “positive regulation of sterol” and “cholesterol transport”, “fascia adherens”, “postsynaptic specialization”, and “asymmetric synapse” pathways. Positive regulation of sterol and cholesterol is crucial for cellular homeostasis, especially lipid metabolism ^52^. Disturbed cholesterol homeostasis in neuronal cells is observed in AD, contributing to tau-related pathogenesis ^53^. Disruptions in postsynaptic specialization may lead to synaptic dysfunction in AD ^54^. Asymmetric synapses, or excitatory synapses, are activated by neurotransmitter release ^55^. Aβ co-localizes with postsynaptic densities, contributing to the loss of excitatory synapses in AD ^56^, suggesting tau-associated gene pathways likely contribute to synaptic dysfunctions.

AD PRS was strongly associated with amyloid-SUVR compared to tau, indicating that AD-associated genetic loci are linked strongly to Aβ pathology but not tau. Amyloid-SUVR PRS was significantly associated with clinical AD, whereas tau PRS showed no association, suggesting different genetic pathways for Aβ and tau in AD pathology. Among *APOE* ε4 carriers, Aβ seems crucial in early AD stages, while tau plays a role later in the middle and late stages of the disease. Hippocampal sclerosis, likely mediated by tau, occurs specifically among *APOE* non-ε4 carriers, though its association with tau PRS showed only a trend towards significance.

Cortical atrophy was associated with rs78636169 in *JARID2* in both ADNI and A4 cohorts. ADNI results showed rs78636169-A allele mediated cortical atrophy in Braak stage 1-6 regions^57^. Conversely, pre-clinical AD participants in the A4 cohort exhibited cortical atrophy in Braak stage 1, representing early tau accumulation stage. Previous studies demonstrated tau accumulation following Braak stages ^58^, and share a similar topography with cortical atrophy patterns ^59^. Consistent with those findings, our results suggest that rs78636169-mediated tau accumulation triggers cortical atrophy in the same topographical regions.

Despite our limited sample size, results obtained through a meta-analysis of two cohorts, enhance the generalizability of our findings due to consistent associations. We identified *JARID2* in the *PRC2* multi-protein complex and *CTNNA3* genes as associated with tau deposition using tau-PET SUVR measures. MR analysis confirmed the role of the PRC2 complex in tau pathology. Brain cortical analysis revealed *JARID2*-mediated atrophy following Braak stages. We also found *COL5A2* and two major pathways involved in amyloid and tau pathologies.

Taken together, the findings here offer valuable insights into tau pathology mediated by the PRC2 complex in AD, paving the way for new therapeutic interventions and understanding genetic loci associated with amyloid and tau in AD progression.

## Data Availability

All data produced are available online at
http://adni.loni.usc.edu
https://www.ukbiobank.ac.uk/

## Acknowledgments

^¶^Data used in preparation of this article were obtained from the Alzheimer’s Disease Neuroimaging Initiative (ADNI) database (adni.loni.usc.edu). As such, the investigators within the ADNI contributed to the design and implementation of ADNI and/or provided data but did not participate in analysis or writing of this report. A complete listing of ADNI investigators can be found at: http://adni.loni.usc.edu/wp-content/uploads/how_to_apply/ADNI_Acknowledgement_List.pdf

Data analysis was supported by grant from the National Institute on Aging U01AG066752 to Dr.Vardarajan. ADNI data collection and sharing for this project was funded by the Alzheimer’s Disease Neuroimaging Initiative (ADNI) (National Institutes of Health Grant U01 AG024904) and DOD ADNI (Department of Defense award number W81XWH-12-2-0012). ADNI is funded by the National Institute on Aging, the National Institute of Biomedical Imaging and Bioengineering, and through generous contributions from the following: AbbVie, Alzheimer’s Association; Alzheimer’s Drug Discovery Foundation; Araclon Biotech; BioClinica, Inc.; Biogen; Bristol-Myers Squibb Company; CereSpir, Inc.; Cogstate; Eisai Inc.; Elan Pharmaceuticals, Inc.; Eli Lilly and Company; EuroImmun; F. Hoffmann-La Roche Ltd and its affiliated company Genentech, Inc.; Fujirebio; GE Healthcare; IXICO Ltd.; Janssen Alzheimer Immunotherapy Research & Development, LLC.; Johnson & Johnson Pharmaceutical Research & Development LLC.; Lumosity; Lundbeck; Merck & Co., Inc.; Meso Scale Diagnostics, LLC.; NeuroRx Research; Neurotrack Technologies; Novartis Pharmaceuticals Corporation; Pfizer Inc.; Piramal Imaging; Servier; Takeda Pharmaceutical Company; and Transition Therapeutics. The Canadian Institutes of Health Research is providing funds to support ADNI clinical sites in Canada. Private sector contributions are facilitated by the Foundation for the National Institutes of Health (www.fnih.org). The grantee organization is the Northern California Institute for Research and Education, and the study is coordinated by the Alzheimer’s Therapeutic Research Institute at the University of Southern California. ADNI data are disseminated by the Laboratory for Neuro Imaging at the University of Southern California.

The A4 Study is a secondary prevention trial in preclinical Alzheimer’s disease, aiming to slow cognitive decline associated with brain amyloid accumulation in clinically normal older individuals. The A4 Study is funded by a public-private-philanthropic partnership, including funding from the National Institutes of Health-National Institute on Aging, Eli Lilly and Company, Alzheimer’s Association, Accelerating Medicines Partnership, GHR Foundation, an anonymous foundation and additional private donors, with in-kind support from Avid and Cogstate. The companion observational Longitudinal Evaluation of Amyloid Risk and Neurodegeneration (LEARN) Study is funded by the Alzheimer’s Association and GHR Foundation. The A4 and LEARN Studies are led by Dr. Reisa Sperling at Brigham and Women’s Hospital, Harvard Medical School and Dr. Paul Aisen at the Alzheimer’s Therapeutic Research Institute (ATRI), University of Southern California. The A4 and LEARN Studies are coordinated by ATRI at the University of Southern California, and the data are made available through the Laboratory for Neuro Imaging at the University of Southern California. The participants screening for the A4 Study provided permission to share their de-identified data in order to advance the quest to find a successful treatment for Alzheimer’s disease. We would like to acknowledge the dedication of all the participants, the site personnel, and all of the partnership team members who continue to make the A4 and LEARN Studies possible. The complete A4 Study Team list is available on: www.actcinfo.org/a4-study-team-lists.

The replication data from the Mayo Clinic contained in this analysis were obtained under research grant U01 AG006786 (PI: Ronald C. Petersen, M.D., Ph.D.) from the National Institutes of Health to the Mayo Clinic Study of Aging.

