## Supplemental Materials for "Genome-wide scan of Flortaucipir PET levels finds *JARID2* associated with cerebral tau deposition"

**Genome-wide scan in Flortaucipir PET studies identifies common variants in the transcriptional repressor *JARID2* associated with Cerebral Tau Deposition**

**Supplementary Methods**

**SNP genotyping and imputation**

The genotype data from A4, ADNI-1, ADNI-GO, ADNI-2, ADNI-3 and ADNI WGS cohorts were downloaded from the ADNI database (<http://adni.loni.usc.edu>). Genotyping methods for these cohorts are available at <https://ida.loni.usc.edu/>. Standard quality control (QC) procedures were performed separately for each genotype dataset. SNPs were excluded if there was a low genotype call rate <95%, Hardy-Weinberg equilibrium (HWE) test *P*-value <1.00×10^-6^ and the minor allele frequency (MAF) <1%. Individuals with low genotype call rate <95%, cryptic relatedness (PI_HAT>0.2) and sex discordance were removed. The number of SNPs and individuals included after quality control is shown in **Supplementary Table 1**. The genotype quality control for A4, ADNI-1, ADNI-GO, ADNI-2 and ADNI-WGS study cohorts are provided in our previous study [1]. ADNI-3 participants were genotyped using the Illumina Infinium Global Screening Array v2 platform. Before genotype QC, ADNI-3 included 327 participants and 759,993 SNPs. After QC, 322 participants and 503,036 SNPs remained.

After quality control, imputation was performed for all study datasets using the TOPMed reference panel implemented in the TOPMed imputation server. After imputation, SNPs with low quality (info score <0.9) and MAF<0.01 were removed. The total number of SNPs remaining after imputation quality control in each study dataset is provided in **Supplementary Table 1**. We included individuals only if they had both genotype data and tau imaging data. This resulted in the inclusion of 733 individuals, which is shown in **Table 1** and 7,725,775 SNPs for the association analysis. Of the 733 mutli-ethnic individuals, 95.8% were non-Hispanic whites, 3.5% were Hispanic, and the remaining 0.7% had unknown ethnicity. To address the population substructure within the dataset, we computed principal components (PCs) from the GWAS data using the “smartpca” script from the EIGENSOFT package. No PCs were significantly associated with tau-SUVRs, so we included the first 3PCs for our analysis.

**Mendelian Randomization analysis**

Instrument selection and MR analysis

To obtain genetic instrumental variables (IVs) for the 84 circulating plasma proteins [2], we extracted *cis*-acting biallelic IVs located within +/- 1Mb of the corresponding gene encoding the protein (defined as *cis*-pQTL [protein quantitative trait loci]). IVs were further filtered by the strength of association (*P*<5×10^-8^), MAF>0.01, and clumped at a pairwise linkage disequilibrium (LD) threshold of r^2^<0.001 in a window of 10000 KB using the TwoSampleMR R package to extract independent genetic variants [3]. IVs with F-statistics <10 was excluded to avoid weak instrument bias [4]. Data was harmonized using the “harmonized” function as implemented in the TwoSampleMR package. We utilized a randomly selected 10,000 European participants from the UKB for extracting IVs and the European reference panel of individuals from the 1000 genomes for performing clumping [5, 6]. We considered Wald ratio (IV<2), or inverse variance weighted (IVW) method (IV ≥ 2) as our primary methods, and IVW method was followed by weighted median and MR-Egger where possible. An FDR-corrected *P*<0.05 was defined as statistically significant.

Co-localization analyses

To avoid confounding by LD in MR, we performed Bayesian genetic co-localization to identify shared causal variants between circulating plasma proteins and tau PET levels in the corresponding genomic region. The co-localization analysis was performed on the pre-defined *cis*-region (i.e., within +/- 500KB) of the corresponding coding gene. Priors were set as default that any SNP within the co-localization window was exclusively associated with the two traits with the probability of 1 × 10^-4^ and associated with both traits with the probability of 1 × 10^-5^ [7]. A co-localization posterior probability (PP) higher than 70% [PPH4>70%] was considered strong evidence that the two traits are likely to co-localize in the region, while 50% < PPH4 < 70% was considered suggestive evidence. The co-localization analysis was conducted using the ‘coloc’ R package [7].

**Gene pathway analysis**

Genes that showed nominally significant (*P* <0.05) with both tau and amyloid were chosen for the functional gene pathway analysis. This gene enrichment analysis used databases such as the Kyoto Encyclopedia of Genes and Genomes (KEGG), Reactome, and Gene Ontology (GO), encompassing information related to cellular composition, molecular function, and biological processes. The pathway analysis was conducted using the R package clusterProfiler, version 4.8.3 [8].

**Polygenic risk score (PRS) analysis**

AD PRS was calculated in the ADNI and A4 datasets using summary statistics obtained from a large-scale AD GWAS conducted by Kunkle et al. in 2019 [9], which included data from 94,437 participants. PRS was estimated using the PRSice version 2 and PLINK software using 152,768 SNPs with a significance level of *P*<0.01 from the AD GWAS. Independent SNPs were computed using linkage disequilibrium (LD) clumping with LD r^2^>0.25. To exclude the effect of the *APOE* gene on the AD-PRS, we excluded SNPs in the 500KB base-pair region flanking the APOE-ε4 rs429358 SNP. Subsequently, we analyzed the relationship between the AD PRS and tau and amyloid-SUVR measures using a linear regression model while controlling for age, sex, and the first 3 PCs. We combined 528 A4 and ADNI participants with both tau and amyloid measurements.

Next, we explored the association of the PRS of tau and amyloid SUVR with clinical and pathological AD status among autopsied patients.

The tau-SUVR PRS was calculated in the Religious Orders Study and the Memory and Aging Project (ROSMAP) dataset using the summary statistics from the A4 and ADNI study cohorts, while the amyloid PRS was estimated using summary statistics from our previous large-scale amyloid GWAS [1]. Detailed descriptions of the ROSMAP study cohort can be found elsewhere [10-12], and demographic characteristics of the study participants are provided in **Supplementary Table 2**. Outcomes included clinical AD diagnosis and NIA-Reagan Institute criteria for the neuropathological diagnosis of AD. We also assessed the association of tau and amyloid PRS with other neuropathological conditions linked to AD, including hippocampal sclerosis. The descriptions for ROSMAP phenotypes are provided elsewhere [13].

B)

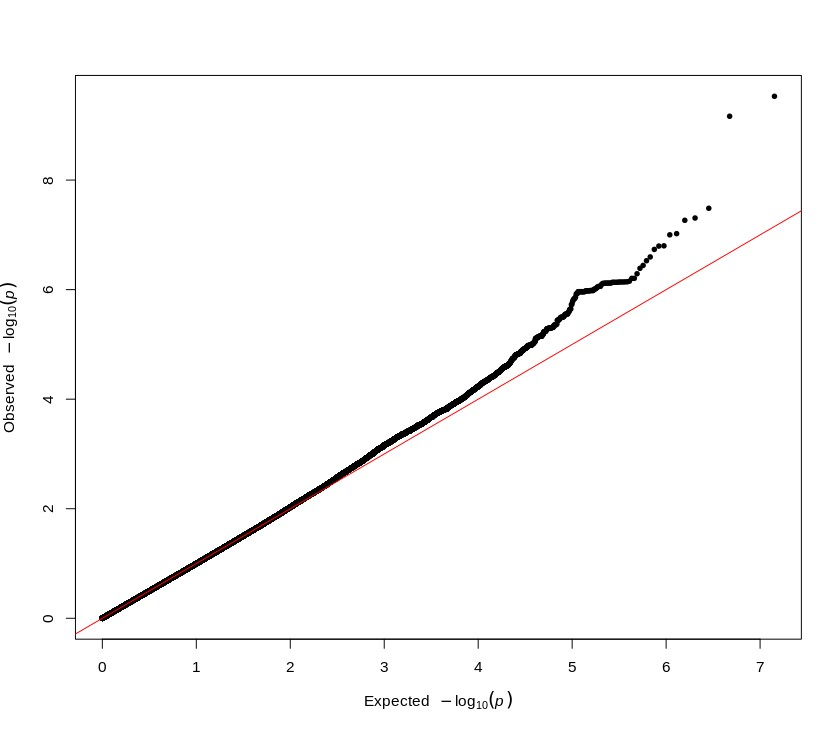

Expected –log10(*p*)

Lambda = 1.001

Observed –log10(*p*)

A)

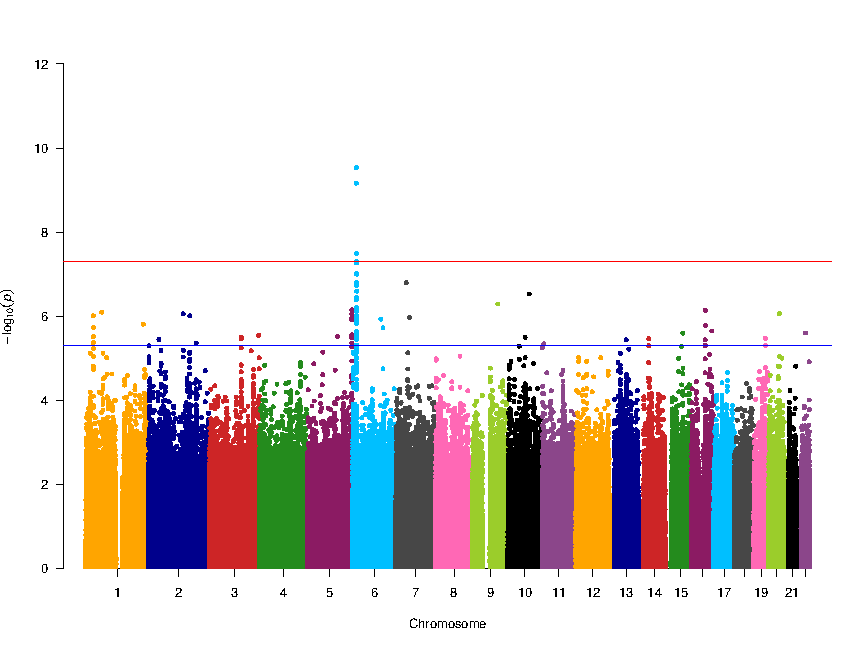

rs78636169

(*JARID2*)

16

18

20

22

Chromosome

-log10(*p*)

**Supplementary Figure 1. SNPs associated with cerebral tau deposition in multi-ethnic subjects. (A)** Manhattan plot showing meta-analysis *P*-values (depicted on the –log_10_ scale) from linear regression on cerebral tau deposition involving multi-ethnic subjects. The A4 cohort was adjusted for age, sex, and three principal components (PCs) for population substructure as covariates, and the ADNI cohort was adjusted for age, sex, diagnosis, and three PCs. The threshold for genome-wide significance is represented by a blue line at *P* = 5×10^-8^, while suggestive significance is indicated by a blue line at *P =* 5×10^-6^ threshold. **(B)** Quantile-Quantile (QQ) plots for the SNPs associated with cerebral tau deposition involving multi-ethnic subjects. The QQ plot showed no spurious genomic inflation (λ = 1.001).

B)

A)

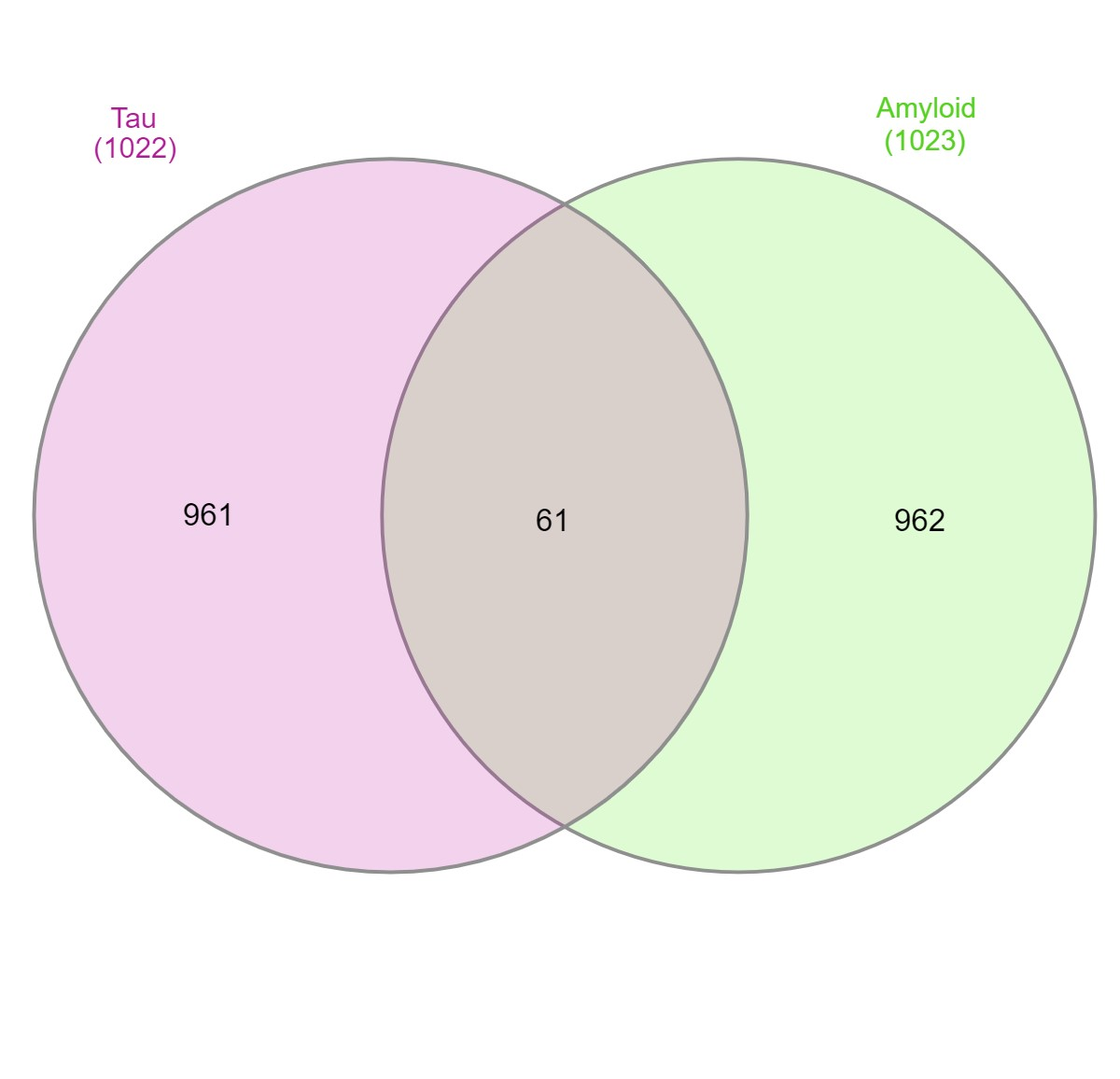

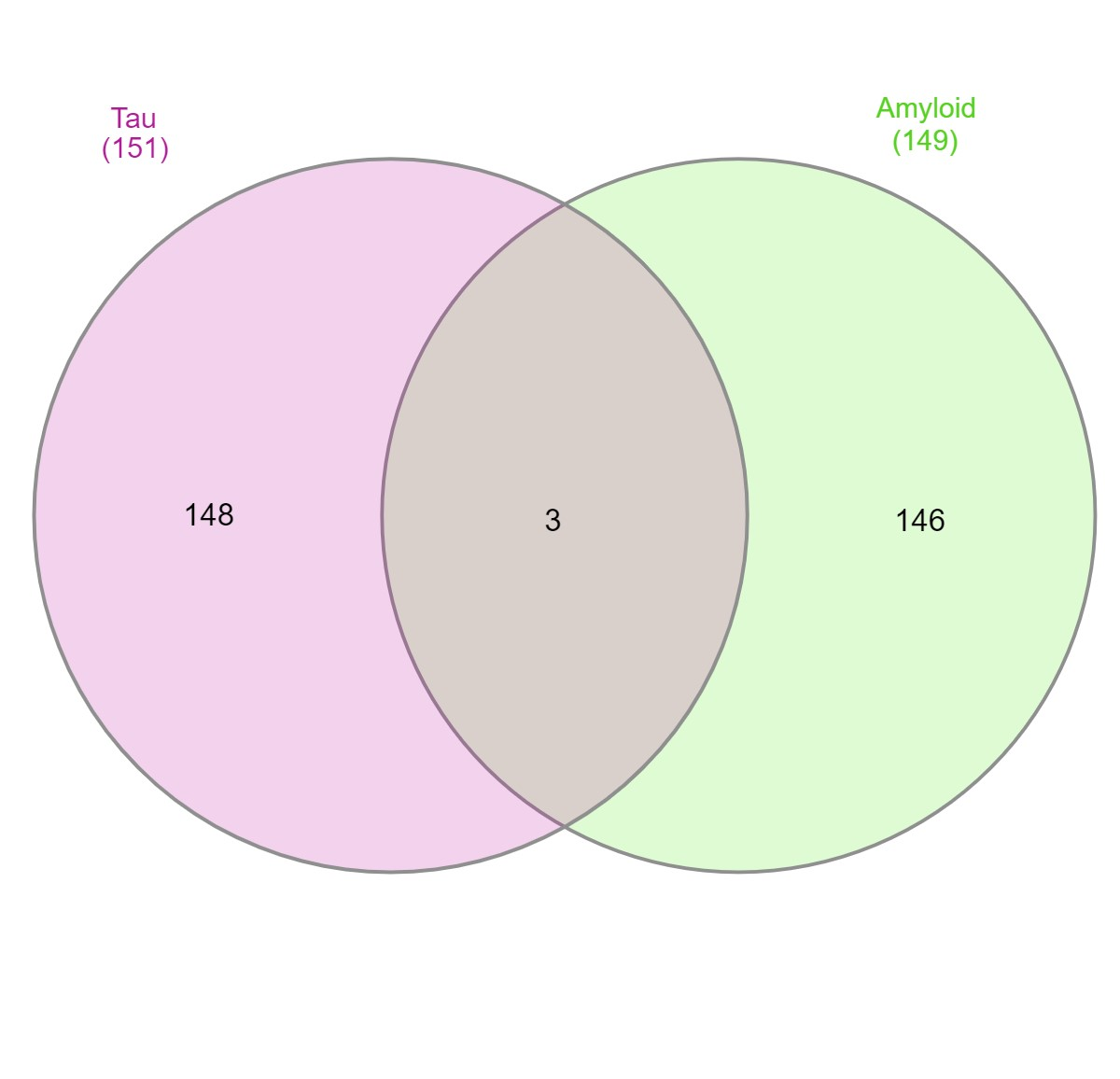

**61 genes with p<0.05 were found**

**to be associated with both tau and amyloid**

**3 genes with p<0.005 were found**

**to be associated with both tau and amyloid**

**(*APOE, TOMM40, COL5A2*)**

**Supplementary Figure 2. Genes associated with amyloid and tau pathologies**. **(A)** Genes that were strongly associated p<0.005 with tau and amyloid deposition were shown in figure **(A)** and the genes with nominal association were shown in figure **(B).**

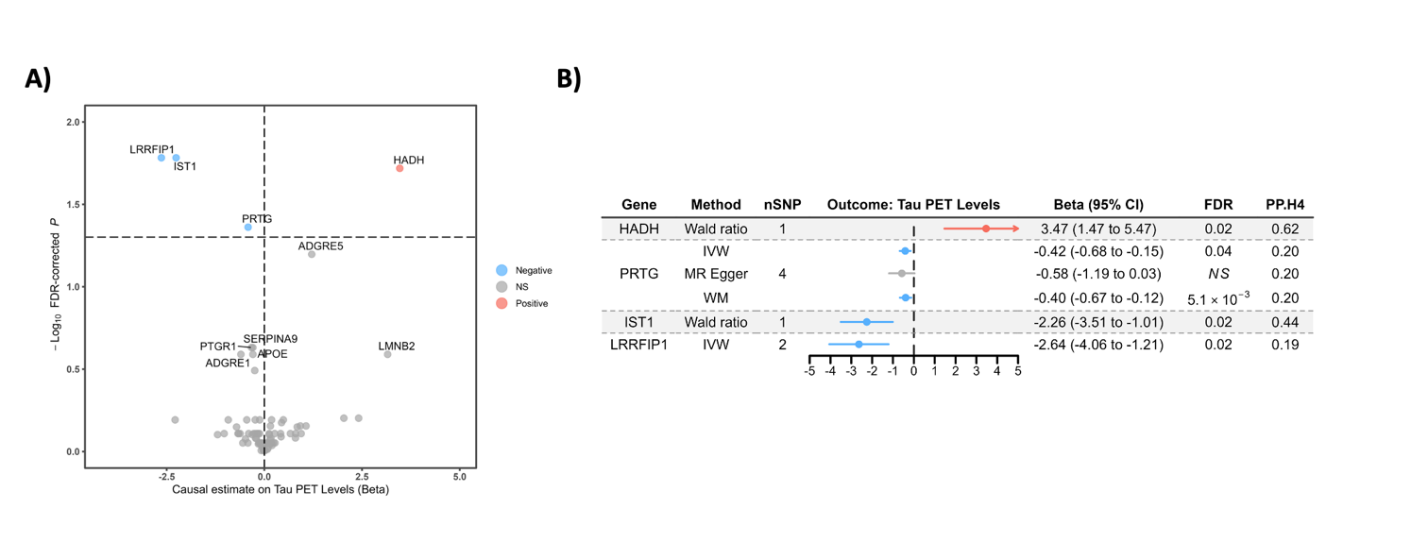

**Supplementary Figure 3. Mendelian Randomization.** MR effect of circulating plasma proteins on PET Tau levels using the UKBPPP proteomics data. **(A)** Volcano plot for the effect of 84 plasma proteins on PET Tau levels and **(B)** Forest plot showing the effect estimates for all proteins surpassing 5% FDR. FDR; false discovery rate, CI; confidence intervals, PP; posterior probability, SNP; single nucleotide polymorphism.

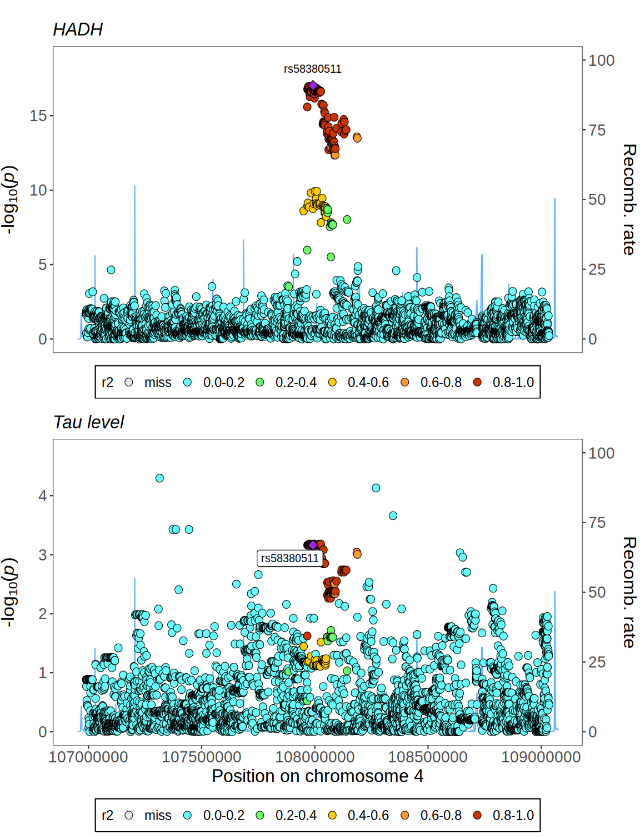

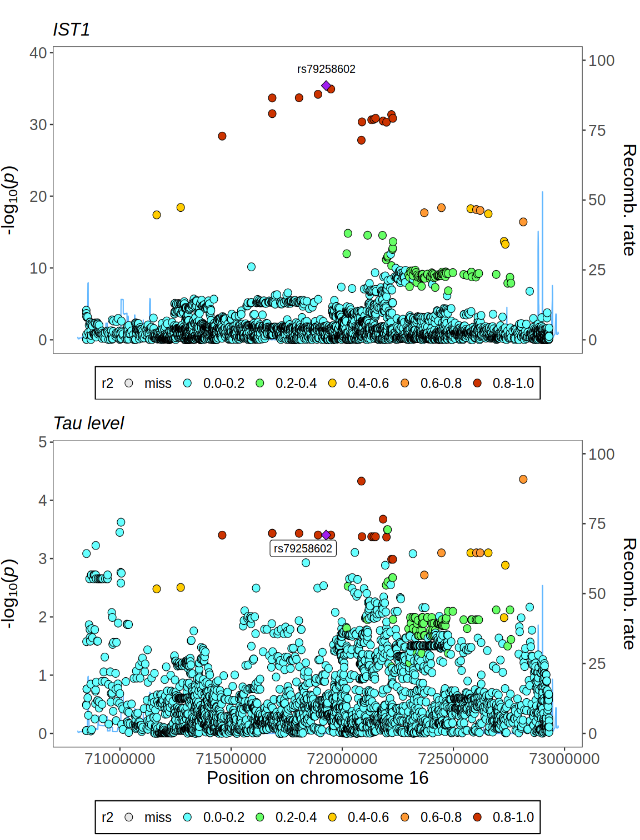

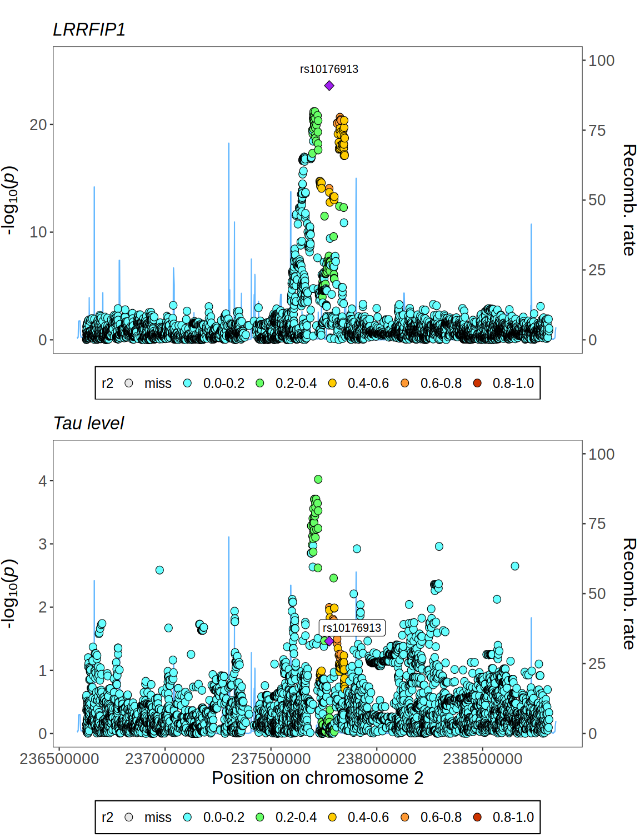

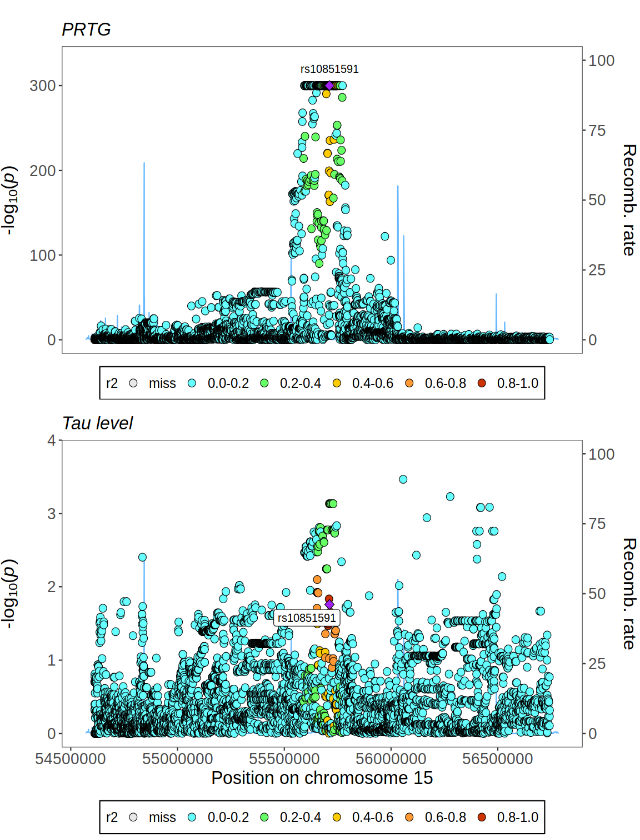

A)

B)

C)

D)

**Supplementary Figure 4. Proteins co-localized with tau levels in Mendelian Randomization**. Co-localization LocusZoom plots showing evidence of genetic co-localization for the **(A)** HADH **(B)** IST1 **(C)** LRRFIP1 and **(D)** PRTG that surpassed a 5% FDR threshold in the MR analysis.

**GO Cellular component pathway**

**GO Biological process pathway**

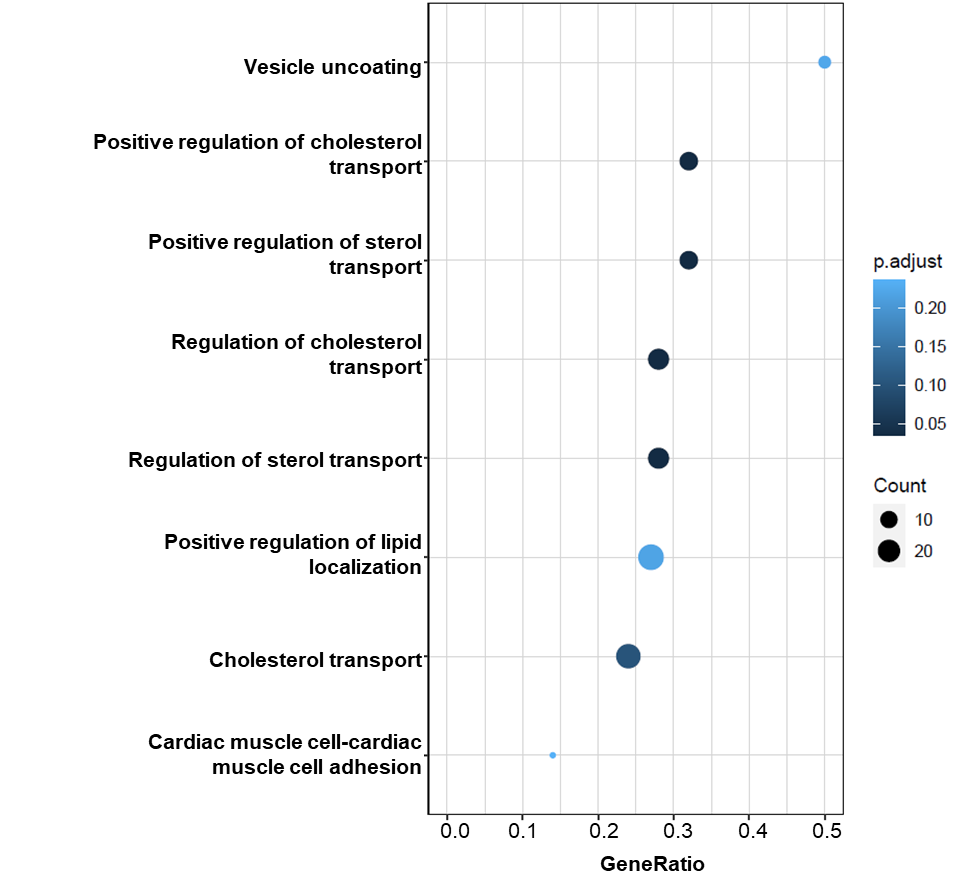

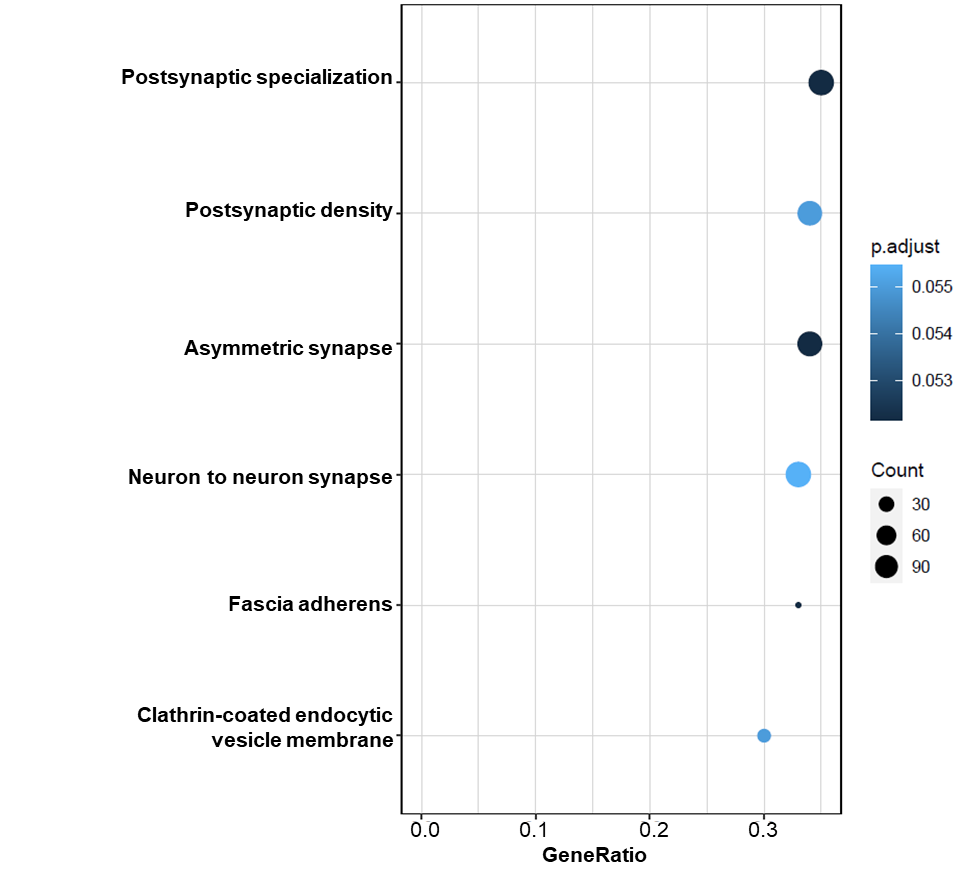

**Supplementary Figure 5. Tau associated (*P*<0.05) genes enriched in Gene Ontology (GO) pathways**. **(A)** GO Biological process pathway **(B)** GO Cellular component pathway.

**GO Cellular component pathway**

**GO Biological process pathway**

**Supplementary Figure 6. Tau and Amyloid associated (*P*<0.05) genes enriched in Gene Ontology (GO) pathways**. **(A)** GO Biological process pathway **(B)** GO Cellular component pathway.

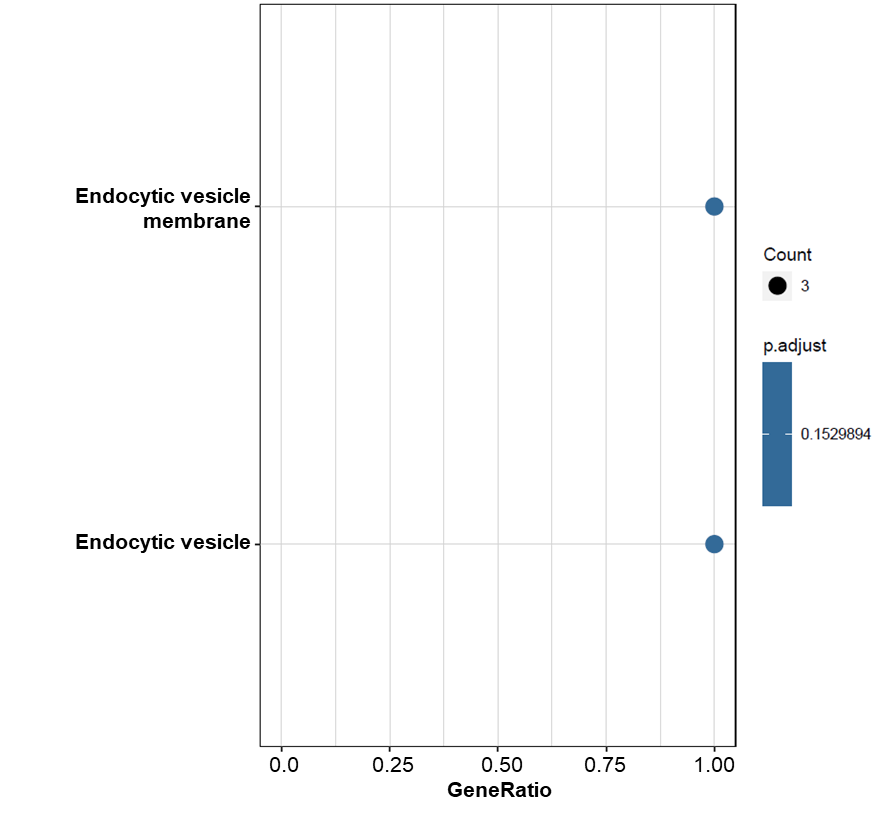

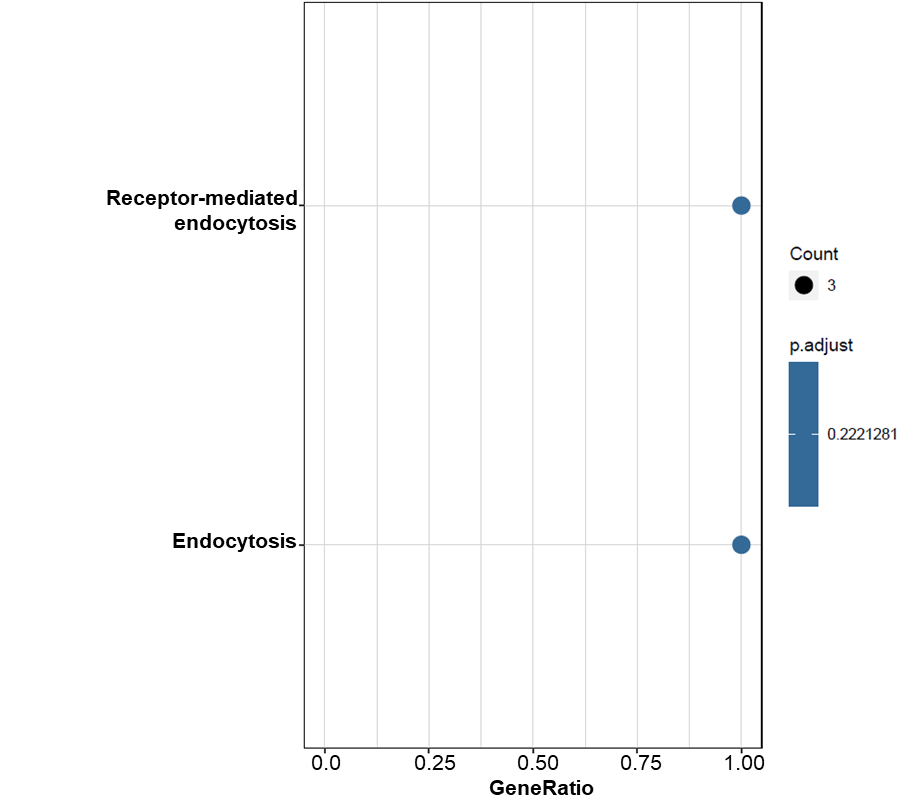
